# Economic evaluation of hospital-to-home transition interventions: a rapid review protocol

**DOI:** 10.1101/2025.05.06.25327087

**Authors:** Evan MacEachern, Michelle L.A. Nelson, Jaylyn Leighton, Marianne Saragosa, Ian D. Graham, Kristina Kokorelias, Paul Herbert, Brina Ludwig-Prout, Kednapa Thavorn

## Abstract

**Background:** Transitions from hospital to home represent a critical juncture associated with hospital readmissions and increased healthcare costs. While interventions exist to improve care transitions, the economic value of these interventions remains limited. By synthesizing economic evaluations of transition interventions, healthcare systems can identify cost-effective strategies to reduce costs and readmission rates to improve outcomes for patients and providers.

**Objectives:** 1) Identify the types of economic analyses commonly applied in hospital-to-home evaluations; 2) summarize the financial implications and economic value of these interventions; 3) compare the costs and cost-effectiveness of interventions across care settings and patient populations.

**Methods:** We will conduct a rapid review to summarize existing literature on the cost-effectiveness of interventions aimed at improving hospital-to-home transitions. We will search electronic databases (Embase, MEDLINE, CINAHL) to identify studies assessing the economic impact of transition interventions on patient and health system outcomes. We will focus on economic studies, including cost studies, cost-consequence analyses, cost-minimization analyses, cost-effectiveness analyses, cost-benefit analyses, or cost-utility analyses, to understand the economic value of these interventions for both healthcare systems and patients.

**Implication:** Evidence on the economic benefits of hospital-to-home interventions can guide efficient and sustainable resource allocation in healthcare systems. Investing in sustainable transition interventions has the potential to lower overall healthcare expenditures while enhancing patient satisfaction and improving health outcomes. We hypothesize that this review will reveal that targeted strategies, such as enhanced discharge planning and home-based discharge supports, can achieve significant cost savings by reducing hospital readmissions and improving patient adherence to post-discharge care plans.

## Introduction

Hospital systems continually aim to enhance healthcare services while managing costs effectively for both patients and health systems.^1,2^ Embedded within the health care system are crucial yet fragmented transitional care pathways from hospital to home.^3–11^ Hospital-to-home transitions involve multiple stages of care, including primary, acute, step-down, or outpatient care, that are essential to rehabilitation but are often perceived as unfamiliar and challenging for patients.^7,11^ To ease the transition from hospital to home, targeted interventions may include comprehensive discharge planning, patient education, follow-up care, telehealth services, or even home visits by healthcare professionals.^11–13^ Transitional interventions such as these can provide individuals with supports and services to manage and cope in their homes and communities safely,^11–13^ which also lowers healthcare expenditures.^1,7,8,11,13^

Economic evaluation of care transition interventions is crucial for fostering effective healthcare teams and enabling stakeholders, including care providers, policymakers, and decision-makers, to make evidence-informed decisions about resource allocation and cost-effective practice.^14^ Understanding the economic implications of hospital-to-home transitions can help prioritize interventions that provide the best economic value and optimize limited healthcare resources. By extension, economic evaluations may also determine if a novel intervention is economically viable for scaling and spreading across various health systems and patient populations.^15^ Existing literature on the economic evaluations of hospital-to-home interventions is emerging. Still, it indicates that these interventions are associated with reduced costs for health systems and services compared to interventions lacking transitional care support.^5,6,13^ However, the absence of a formal knowledge synthesis limits our understanding of the economic utility of hospital-to-home interventions. Without a comprehensive overview of economically viable practices during transitional care points, there is a risk of misallocating human and fiscal resources and misinterpreting the perceived benefits of these interventions.

We aim to summarize the methodologies used in these economic evaluations to inform the design of an economic evaluation for a recent CIHR-funded implementation-effectiveness randomized controlled trial (FRN: 191308). This approach will enable us to build on existing successes and learn from past shortcomings rather than starting anew. Accordingly, this manuscript describes the protocol for a rapid review that outlines our inquiry into the economic value of hospital-to-home transition interventions, focusing on the economic analyses used to inform health care system decision-making.

## Objectives

This paper aims to present a detailed protocol for a rapid review aimed at synthesizing existing evidence on economic analyses of hospital-to-home transition interventions. Specifically, we will: 1) identify the range of dominant approaches used in economic analyses of hospital-to-home evaluations; 2) summarize the financial implications and economic value of these interventions as reported by included articles; and 3) compare the costs and cost-effectiveness of interventions across care settings and patient populations. In doing so, we will also identify gaps in the current evidence base and suggest future research on economic evaluations of hospital-to-home transition interventions.

## Materials and methods

The present paper is a protocol for a rapid review. Described as “within the family” of systematic reviews,^16^ rapid reviews offer a methodical and comprehensive approach to evaluating an existing body of research on a specific topic. They differ from traditional systematic reviews because they prioritize speed and efficiency. Rapid reviews are designed to quickly identify and summarize relevant literature. Previous literature supports the effectiveness of rapid reviews in informing health policy and decision-making.^17^ Accordingly, this rapid review protocol follows Tricco et al.’s (2017)16 guidance and serves as a foundation for a broader research agenda examining hospital-to-home care transitions. We will begin this review in July 2025 and anticipate completing it within 6 months by December 2025. We intend to register this review with PROSPERO upon initiation of the review.

### Participant and public involvement

Individuals with lived experience have been integral members of our larger research team. While they were not directly engaged in designing the methods for this protocol, their insights were instrumental in identifying the need for this rapid review and shaping our plan for a primary economic evaluation of hospital-to-home interventions, as they contributed to the development of our grant proposal. To ensure the relevancy of this work, we have aligned the goals of this rapid review with the perspectives of team members with lived experience, who will also play a key role in interpreting the results of this review.

### Stage 1: Identifying the research questions (RQ)

The purpose of this review is to consolidate existing literature on economic evaluations of hospital-to-home transition interventions. Using a systematic methodological approach, this review seeks to address the following research questions (RQs):

> RQ1) Which types of economic analyses are used to evaluate hospital-to-home transition interventions?
>
> RQ2) What are the costs and cost-effectiveness of hospital-to-home transition interventions?
>
> RQ3) How do the costs or cost-effectiveness of these interventions vary based on the care setting (e.g., primary care, acute care, rehabilitation, outpatient and community reintegration) or the patient population being studied?

### Stage 2: Identifying relevant literature

To identify relevant peer-reviewed literature, a comprehensive search will be conducted across the following electronic databases: MEDLINE (Ovid), EMBASE (Elsevier), and CINAHL. The electronic database search will be supplemented by searching PROSPERO and the Cochrane Reviews Library for relevant reviews. Our search strategy will be vetted by an experienced Information Scientist at Sinai Health System, Toronto, ON, and will be uploaded for peer review using Peer Review of Electronic Search Strategies (PRESS) 2015 guidelines.^18^ We will also scan the reference lists of extracted articles to identify additional relevant literature that our search strategy may have missed. Subject headings and text related to the concepts of interest will be incorporated into the search: “hospital-to-home transition interventions” and “economic studies.” We define “hospital-to-home transition interventions” as strategies designed to facilitate a smooth transfer of patients from hospital discharge to home settings, including various clinical and supportive measures to improve post-discharge outcomes and reduce healthcare costs. We will include economic studies, encompassing cost studies, cost-consequence analysis, cost-minimization, cost-benefit analysis, cost-effectiveness analysis, and cost-utility analysis. Articles included in our rapid review must be published in English and involve human participants aged 18 and older. To meet the inclusion criteria, all included articles must comprehensively describe the hospital-to-home intervention. To ensure the literature remains relevant to the scope of our review, we will include peer-reviewed articles, grey literature, magazine articles, and conference abstracts. As our review has a broad inclusion of study designs, all peer-reviewed academic literature will be assessed for risk of bias using the Mixed Methods Appraisal Tool (MMAT) Version 2018.^19^ Risk of bias will be evaluated by two reviewers as advised by MMAT authors.^19^ Appendix A provides an example of the search strategy we implemented in Medline (date October 29th, 2024).

### Stage 3: Study selection

Study selection will follow a two-step, multi-reviewer screening process, beginning with a title and abstract screening followed by a full-text review. We will consider all primary studies, including experimental, observational, and mixed methods designs. We will include any adult (18 years or older) participant population. Publications listed as reviews (e.g., systematic, scoping, and rapid reviews) will be excluded. However, we will examine the reference lists of these articles to identify relevant literature. Before the initial screening, reviewers will conduct a screening exercise with a random set of 25-50 titles and abstracts to ensure reviewer consistency (inter-rater agreement) and to identify articles that meet our rapid review inclusion criteria. At this time, reviewers will discuss the results of the training exercise to ensure congruity of article selection. The reviewers will make any necessary revisions to clarify the inclusion criteria and ensure alignment with their review process.^20^

The finalized rapid review search will be exported to Covidence^21^ to remove duplicates and screen for study inclusion. Then, reviewers will independently evaluate the titles and abstracts per our review’s eligibility criteria, categorizing articles into “yes,” “no,” and “maybe” distinctions. An interrater reliability statistic will be calculated according to Cohens kappa, a common measure for reviews of two raters.^22^ Reviewers will review all full texts provided a “yes” distinction upon the title and abstract screening. Articles deemed “maybe” will be discussed by reviewers at consensus meetings for final inclusion. Any disagreements or conflicts will be resolved through team discussions with reviewers M.L.A.N and K.T. The full text of articles meeting our eligibility criteria will be retrieved, with citation details imported into Zotero referencing software.^23^ Following the Preferred Reporting Items for Systematic Reviews and Meta-Analyses (PRISMA) guidelines,^24^ we will include a figure detailing the reasons for full-text exclusion, as illustrated in Fig 1.

**Figure 1. Example PRISMA flow diagram of rapid review search results**.

### Stage 4: Data extraction, analysis, & interpretation

Two independent reviewers will extract data from all included articles in duplicate and verify each other’s extraction to ensure reliability of the extracted data. We will utilize a team-developed pilot data extraction forms to gather data relevant to the proposed RQs (see S1 Table, S2 Table). Extracted data will include author details, year of publication, overarching purpose of study, study population, hospital care setting, country in which the study was conducted, type of economic studies, approach of economic evaluation (person-level vs. model-based), time horizon, primary outcomes of interest, program details of hospital-to-home intervention, study results. Before starting data extraction, reviewers. will test the pilot data extraction form on five (5) articles and, if needed, revise the form to ensure it captures sufficient information.^18^ Disagreements that may arise will be resolved through consultation with reviewers M.L.A.N. and K.T. All data generated from this review will be managed and stored on a password-secured hospital network prior to publishing the results as publicly available information.

### Stage 5: Summary and reporting of findings

We will narratively summarize the results and provide an overview of the economic evaluations employed in hospital-to-home transitional interventions. We will report on the frequency of varying types of economic evaluations found in the literature, the settings and contexts in which they are conducted, the participant populations involved, the details of the hospital-to-home transition interventions, and the types of articles included. This information will be presented in tables and figures, accompanied by a narrative description of the findings. Since the reporting guidelines for rapid reviews are still under development, we will adhere to the PRISMA Extension for Scoping Reviews Checklist.^16^ We will report the approach for included interventions using the Template for Intervention Description and Replication (TIDiER) Checklist.^26^

### Phase 6: Ethics, knowledge dissemination, & translation

This rapid review protocol does not require ethics approval. All information and data collected for inclusion in the rapid review will be publicly available. We plan to disseminate the findings through publications and presentations by our research group. We anticipate producing up to two publications from this research: first, the present protocol outlining the intended research and its implications for practice, and second, the results of the rapid review of economic evaluations of hospital-to-home transition interventions. This research will form the foundation for the economic evaluation component of our CIHR-funded trial and will contribute to the development of our own economic evaluation. We anticipate that this work will provide valuable insights into the economic considerations of hospital-to-home transition interventions in post-hospital rehabilitation.

## Data Availability

No datasets were generated or analysed during the current study. All relevant data from this study will be made available upon study completion.

## Author contributions

M.L.A.N. – conceptualization, investigation, methodology, writing, project administration, supervision. E.M. – conceptualization, investigation, methodology, visualization, writing, project administration. J.L. – investigation, writing. M.S. – writing (review and editing), supervision. I.D.G. – conceptualization, writing (review and editing), supervision. K.K. – writing (review and editing), supervision. P.H. – supervision. K.T. - conceptualization, investigation, methodology, writing, supervision.

## Acknowledgements

We are grateful to Andrea Slonosky, MA, who guided the development of our literature search strategy.

## Supporting information

S1 Table. Study and program characteristics of included articles.

S2 Table. Program characteristics of included articles.

S1 File. Search strategy for Medline (Ovid) database.

S2 File. PRISMA-P (Preferred Reporting Items for Systematic review and Meta-Analysis Protocols) 2015 checklist: recommended items to address in a systematic review protocol*

---

**S1 Table.**
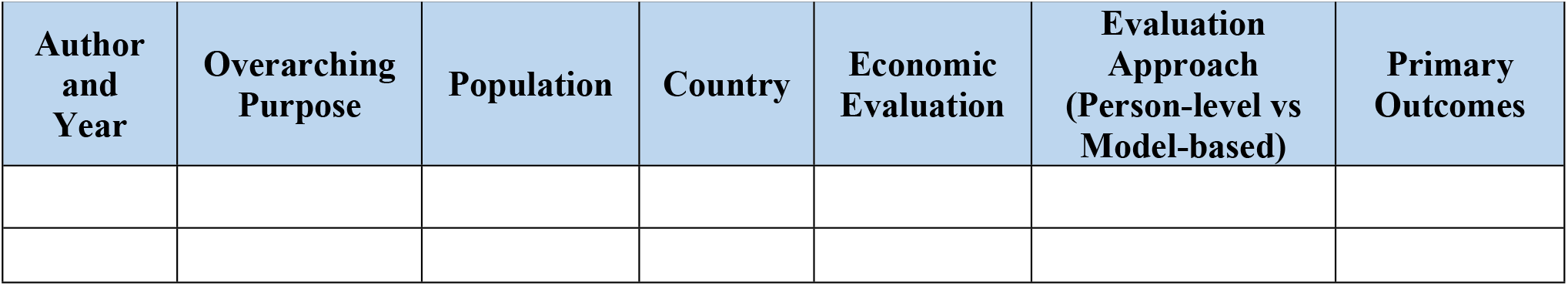
Study characteristics of included articles.

**S2 Table.**
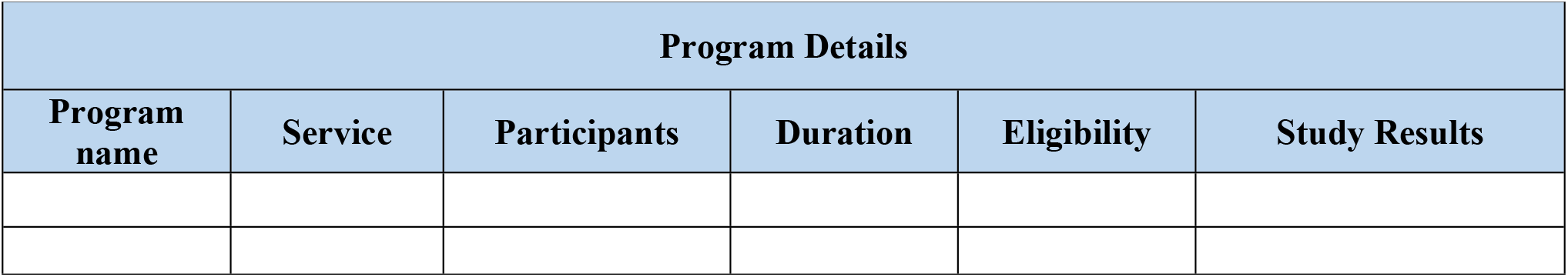
Program characteristics of included articles.

| **Program Details** |  |  |  |  |  |
| --- | --- | --- | --- | --- | --- |
| **Program name** | **Service** | **Participants** | **Duration** | **Eligibility** | **Study Results** |

**S1 File.** Search strategy for Medline (Ovid) database.

| Search # | Query | Results |
| --- | --- | --- |
| 1 | (patient adj1 (release* or discharge)).ti,ab,kf. | 3167 |
| 2 | (hospital adj1 (discharge or release)).ti,ab,kf. | 37856 |
| 3 | exp Patient Discharge/ or Patient Discharge.mp. | 44245 |
| 4 | "Economics".mp. | 512552 |
| 5 | "Costs and Cost Analysis"/ | 52087 |
| 6 | (economic and model*).ti,ab. | 74855 |
| 7 | "cost minimi*".ti,ab. | 1774 |
| 8 | "cost-utilit*".ti,ab. | 6560 |
| 9 | "health utilit*".ti,ab. | 2963 |
| 10 | "economic evaluation*".ti,ab. | 16486 |
| 11 | "economic review*".ti,ab. | 306 |
| 12 | "cost outcome*".ti,ab. | 1047 |
| 13 | "cost analys*".ti,ab. | 9609 |
| 14 | "economic analys*".ti,ab. | 8888 |
| 15 | (budget* and "impact analys*").ti,ab. | 1135 |
| 16 | "cost-effective*".ti. | 36915 |
| 17 | pharmacoeconomic*.ti. | 1886 |
| 18 | "pharmaco-economic*".ti. | 135 |
| 19 | "cost-benefit".ti. | 2612 |
| 20 | costs.ti. | 40444 |
| 21 | "cost-effective*".ot. | 10 |
| 22 | pharmacoeconomic*.ot. | 0 |
| 23 | "pharmaco-economic*".ot. | 0 |
| 24 | "cost-benefit".ot. | 17 |
| 25 | Costs.ot. | 7 |
| 26 | "life year".ti,ab. | 10250 |
| 27 | "life years".ti,ab. | 20606 |
| 28 | galy*.ti,ab. | 126 |
| 29 | "cost-benefit analys*".ti,ab. | 5481 |
| 30 | "cost-effectiveness analys*".ti,ab. | 16128 |
| 31 | (cost or economic*).ti. or (cost or economic*).ot. | 161124 |
| 32 | 4 or 5 or 6 or 7 or 8 or 9 or 10 or 11 or 12 or 13 or 14 or 15 or 16 or 17 or 18 or 19 or 20 or 21 or 22 or 23 or 24 or 25 or 26 or 27 or 28 or 29 or 30 or 31 | 675057 |
| 33 | "costs".ti,ab. | 290453 |
| 34 | "cost-effectiveness".ti,ab. | 81365 |
| 35 | markov.ti,ab. | 30417 |
| 36 | "monte carlo".ti,ab. | 63008 |
| 37 | "model".ti,ab. | 2953877 |
| 38 | ("modeling" or "modelling").ti,ab. | 450893 |
| 39 | 33 or 34 or 35 or 36 or 37 or 38 | 3519856 |
| 40 | 32 and 39 | 228950 |
| 41 | exp Quality Improvement/ or Quality Improvement.mp. | 78778 |
| 42 | (organization adj2 management).ti,ab,kf. | 1458 |
| 43 | exp Program Development/ or Program Development.mp. | 33313 |
| 44 | exp Program Evaluation/ or Program Evaluation.mp. | 92274 |
| 45 | exp Pilot Projects/ or "pilot project".ti,ab,kf. | 157933 |
| 46 | (project or program or scheme or plan or initiative or strategy).ti,ab,kf. | 1873581 |
| 47 | measures taken.ti,ab,kf. | 4064 |
| 48 | plan of action.ti,ab,kf. | 1371 |
| 49 | (pediatric* or adolescen* or infan* or child* or matern* or pregnan* or birth*).ti,ab,kf. | 3275740 |
| 50 | 1 or 3 | 44553 |
| 51 | 41 or 42 or 43 or 44 or 45 or 46 or 47 or 48 | 2103950 |
| 52 | 40 and 50 and 51 | 454 |
| 53 | 52 not 49 | 400 |
| 54 | limit 53 to (english language and humans and yr="2000 - Current") | 336 |

**S2 File.** PRISMA-P (Preferred Reporting Items for Systematic review and Meta-Analysis Protocols) 2015 checklist: recommended items to address in a systematic review protocol*.

| Section and topic | Item No | Checklist item |
| --- | --- | --- |
| **ADMINISTRATIVE INFORMATION** |  |  |
| Title: |  |  |
| Identification | 1a | Identify the report as a protocol of a systematic review |
| Update | 1b | If the protocol is for an update of a previous systematic review, identify as such |
| Registration | 2 | If registered, provide the name of the registry (such as PROSPERO) and registration number |
| Authors: |  |  |
| Contact | 3a | Provide name, institutional affiliation, e-mail address of all protocol authors; provide physical mailing address of corresponding author |
| Contributions | 3b | Describe contributions of protocol authors and identify the guarantor of the review |
| Amendments | 4 | If the protocol represents an amendment of a previously completed or published protocol, identify as such and list changes; otherwise, state plan for documenting important protocol amendments |
| Support: |  |  |
| Sources | 5a | Indicate sources of financial or other support for the review |
| Sponsor | 5b | Provide name for the review funder and/or sponsor |
| Role of sponsor or funder | 5c | Describe roles of funder(s), sponsor(s), and/or institution(s), if any, in developing the protocol |
| **INTRODUCTION** |  |  |
| Rationale | 6 | Describe the rationale for the review in the context of what is already known |
| Objectives | 7 | Provide an explicit statement of the question(s) the review will address with reference to participants, interventions, comparators, and outcomes (PICO) |
| **METHODS** |  |  |
| Eligibility criteria | 8 | Specify the study characteristics (such as PICO, study design, setting, time frame) and report characteristics (such as years considered, language, publication status) to be used as criteria for eligibility for the review |
| Information sources | 9 | Describe all intended information sources (such as electronic databases, contact with study authors, trial registers or other grey literature sources) with planned dates of coverage |
| Search strategy | 10 | Present draft of search strategy to be used for at least one electronic database, including planned limits, such that it could be repeated |
| Study records: |  |  |
| Data management | 11a | Describe the mechanism(s) that will be used to manage records and data throughout the review |
| Selection process | 11b | State the process that will be used for selecting studies (such as two independent reviewers) through each phase of the review (that is, screening, eligibility and inclusion in meta-analysis) |
| Data collection process | 11c | Describe planned method of extracting data from reports (such as piloting forms, done independently, in duplicate), any processes for obtaining and confirming data from investigators |
| Data items | 12 | List and define all variables for which data will be sought (such as PICO items, funding sources), any pre-planned data assumptions and simplifications |
| Outcomes and prioritization | 13 | List and define all outcomes for which data will be sought, including prioritization of main and additional outcomes, with rationale |
| Risk of bias in individual studies | 14 | Describe anticipated methods for assessing risk of bias of individual studies, including whether this will be done at the outcome or study level, or both; state how this information will be used in data synthesis |
| Data synthesis | 15a | Describe criteria under which study data will be quantitatively synthesised |
| | 15b | If data are appropriate for quantitative synthesis, describe planned summary measures, methods of handling data and methods of combining data from studies, including any planned exploration of consistency (such as $I^2$, Kendall's $\tau$) |
|  | 15c | Describe any proposed additional analyses (such as sensitivity or subgroup analyses, meta-regression) |
|  | 15d | If quantitative synthesis is not appropriate, describe the type of summary planned |
| Meta-bias(es) | 16 | Specify any planned assessment of meta-bias(es) (such as publication bias across studies, selective reporting within studies) |
| Confidence in cumulative evidence | 17 | Describe how the strength of the body of evidence will be assessed (such as GRADE) |

**\* It is strongly recommended that this checklist be read in conjunction with the PRISMA-P Explanation and Elaboration (cite when available) for important clarification on the items. Amendments to a review protocol should be tracked and dated. The copyright for PRISMA-P (including checklist) is held by the PRISMA-P Group and is distributed under a Creative Commons Attribution Licence 4.0.**
*From: Shamseer L, Moher D, Clarke M, Ghersi D, Liberati A, Petticrew M, Shekelle P, Stewart L, PRISMA-P Group. Preferred reporting items for systematic review and meta-analysis protocols (PRISMA-P) 2015: elaboration and explanation. BMJ. 2015 Jan 2;349(jan02 1):g7647.*

